# A qualitative exploration of patient flow in a developing Caribbean emergency department

**DOI:** 10.1101/2020.05.29.20110759

**Authors:** Loren De Freitas, Steve Goodacre, Rachel O’Hara, Praveen Thokala, Seetharaman Hariharan

**Affiliations:** School of Health and Related Research (ScHARR), University of Sheffield, 30 Regent Street, Regent Court, Sheffield S1 4DA, UK; Anaesthesia and Intensive Care Unit, University of the West Indies, Eric Williams Medical Science Complex, Champs Fleurs, Trinidad and Tobago

**Author notes:** Corresponding author: Loren De Freitas, School of Health and Related Research (ScHARR), University of Sheffield, 30 Regent Street, Regent Court, Sheffield S1 4DA, UK.

**Keywords:** patient flow, emergency departments, developing countries, Caribbean, qualitative

## Abstract

**Objectives:** Emergency departments (EDs) are complex adaptive systems and improving patient flow requires understanding how ED processes work. This is important for developing countries where flow concerns are compounded by resource constraints. The Caribbean is one region with developing emergency care systems and limited research in the area. This study aimed to explore the patient flow process in an emergency department in Trinidad and Tobago, identifying organizational factors influencing patient flow.

**Methods:** Multiple qualitative methods, including non-participant observations, observational process mapping and informal conversational interviews were used to explore patient flow. The process maps were generated from the observational process mapping. Thematic analysis was used to analyze the data.

**Setting:** The study was conducted at a major tertiary level emergency department in Trinidad and Tobago.

**Participants:** Patient and staff journeys in the emergency department were observed.

**Results:** Six broad categories were identified-1) ED organizational work processes, 2) ED design and layout, 3) material resources, 4) nursing staff levels, roles, skill mix and use 5) non-clinical ED staff and 6) external clinical and non-clinical departments. The study findings were combined with existing literature to produce a model of factors influencing ED patient flow. Barriers and facilitators to patient flow were highlighted.

**Conclusion:** The knowledge gained may be used to strengthen the emergency care system in the local context. The model of ED patient flow may be used to systematically examine factors influencing patient flow, informing policy and practice. However, the study findings should be validated in other settings.

**Article summary:** *Strengths and limitations of this study:* Previous studies have been predominantly conducted in developed countries using quantitative methods Strengthening emergency care systems is becoming a priority in developing countries but the Caribbean remains an under-represented region. This study explores ED patient flow in a developing Caribbean country using a multi-method qualitative design, primarily observational process mapping Single observer used to collect data Singe site may produce context specific findings

## INTRODUCTION

Improving ED patient flow requires understanding the work processes that create flow problems [1]. For this study, ED patient flow has been defined as the progressive movement of patients through care processes, where movement refers to the transformation of an input activity to an output, from arrival until the patient physically leaves the emergency department [2, 3]. Most previous studies addressing ED flow have been conducted in developed settings, focusing on effectiveness of interventions, but have not explored how and why the intervention was (un)able to produce its effect, which is important for generalizability of findings [4].

Implementing interventions without understanding and optimizing factors that influence flow may worsen any inappropriate use of resources, increasing costs, leading to an unproductive system [5]. This is particularly important in developing countries or developing emergency care systems where flow concerns are often compounded by limited resources and a lack of protocols to mitigate issues. In these settings, it is essential to develop robust, effective emergency systems as disease and migration patterns shift, burdening systems [6]. The World Health Organisation (WHO) has placed strengthening emergency care systems on its agenda and consensus statements have noted that emergency care research in developing countries should include ED organization and system design studies [6, 7, 8].

Trinidad and Tobago is a developing country in the Caribbean with a developing emergency care system. The health system is a mix of public and private facilities [9]. One previous study in Trinidad evaluated the usefulness of simulation modeling as a management tool to optimize an ED process [10]. Although the study determined that simulation modeling was a useful tool to identify bottlenecks, a detailed analysis of factors influencing the patient flow process was not presented [10]. Conducting research in developing settings, like the Caribbean, is essential to determine generalizability and transferability of knowledge on patient flow from developed settings as well as gaining new insights from developing settings.

Current literature on ED patient flow has an abundance of quantitative studies but limited qualitative studies exploring the area [4,11]. This study aimed to use qualitative observational methods to identify organisational factors influencing patient flow in an emergency department in Trinidad and Tobago.

## METHODS

### Study design

A pragmatic-critical realist approach was adopted using an exploratory case study design [12,13]. Multiple qualitative methods were used including non-participant observations, observational process mapping and field conversations. These methods were not distinct, independent methods but rather the qualitative process was flexible and iterative with methods overlapping. Observational process mapping utilised direct observations to identify process steps such as activities, delays and decisions as well as what is happening to the patient [14]. Maps reflect the patient process in its current form and are created as patients experience the process and not on perception or assumptions. In process mapping, varying details of the steps in the process may be presented. For example, a low level map will present details of every single step in the process whereas medium and high level maps may only present significant or sustained steps in the process. In this study, medium level maps are presented [14].

### Study setting

The setting was an emergency department in a major public teaching hospital in Trinidad and Tobago which had approximately 450 beds and an estimated 72,000 ED attendances annually. The ED utilised the Canadian Triage and Acuity Scale (CTAS) [15]. ED areas reflected CTAS triage levels with a separate area for minor trauma patients (supplementary file 1 for schematic layout of ED).

### Ethical approval

The University of the West Indies Campus Ethics Committee and the hospital site granted ethical approval (CEC014/09/16).

### Data collection and processing

Data were collected by the lead author, a PhD student familiar with the ED site. The research team consisted of an emergency physician, a qualitative researcher, an health economist and a local researcher. This collaborative approach served to limit the influence of any one researcher’s background on the study. A pilot study was conducted in April 2017 to practice the process mapping technique and uncover any practical issues. Data were then collected from May to August 2017 with a follow-up session in November 2017.

Posters were displayed throughout the ED for the study period. These served to provide information on the study and inform the entire ED population that research was being conducted. When staff and patients were approached, verbal consent was obtained and participants were reminded that they did not have to participate. Purposeful sampling utilised variables such as staff experience, triage category and weekday to develop an in-depth understanding of the patient flow process exploring potential variation amongst triage categories, day of week and crowded periods. Observations were conducted on all seven days of the week and lasted from three to six hours to limit researcher fatigue. In total, the data collection covered a 24-hour period in each of the main ED areas (6am-12pm, 12pm-6pm, 6pm-12am, 12 am-6am). Data collection continued until no new ideas, patterns and themes emerged [16].

In this study, the maps reflected the general organisational ED patient flow process rather than the process for a single patient or a clinical diagnosis/pathway. Steps taken by patients were recorded as they entered an ED area. In areas with high patient turnover (Eg. Triage), the number of ED patient journeys mapped was greater than in the other areas. If a patient was significantly delayed at a step (more than one hour), the researcher then began observing another patient. Observations concluded when the patient’s ED journey was complete or the observation time period ended. Observations focused on activity within the step as well activity around the patient with the aim of understanding how the process worked and why things occurred as they did.

During the study, the department underwent a reconfiguration which was independent of the study. Since the reconfiguration provided an opportunity to observe and map the effects of the changes, the data collection period was extended to incorporate the changes. Detailed handwritten field and reflexive notes were recorded and transcribed into Microsoft Word 2016. Files were anonymised and labelled. Recording verbatim speech was difficult but ‘speech in action’ was included which described actions and speech used by participants as they occurred [17].

Process maps were constructed in Edraw Max 9.4 software. Review of maps occurred over four sessions from February-March 2018. Key staff members validated the maps, providing feedback, clarifying uncertain areas. Staff members included a consultant, head nurse, senior doctor and one representative each from the point of care testing lab, escort services and ED radiology department. Each session lasted approximately one hour. A scribe was present to record the data.

### Patient and Public involvement

This research was not conducted with patient and public involvement as it is not established in Trinidad and Tobago.

### Data analysis

Data were analysed with thematic analysis [18]. NVIVO 11 software facilitated the analysis. Analysis was an iterative process with preliminary analyses starting during fieldwork to allow for data saturation and continued into final analysis and interpretation phases. Codes and themes were inductively generated from the data but were influenced by descriptors developed in comprehensive literature reviews conducted prior to data collection [11]. Thus, while the emphasis was on the generation of data driven codes and themes, if there was a similar descriptor from the literature reviews, it was used. As qualitative research focuses on range and diversity of data, themes were based on relevance to the research question and not on number of occurrences in the data [19]. A selection of transcripts and analytical themes were discussed with the co-authors who provided critical feedback.

## RESULTS

A total of 203 hours of observations were conducted which included 48 hours of nonparticipant observations and 155 hours of observational process mapping with 143 ED patient journeys mapped. Of these, 23 were categorised as CTAS Level 1-3, 32 as Level 4, 21 as Minor Operating Theatre (MOT) and 67 were Registration/Triage/CTAS Level 5 patients.

### Summary of process maps

Four process maps were generated from the observational process mapping (figures 1-4). The main process map (figure 1) represents the ED patient flow process from entry to exit. On arrival to the ED, a triage nurse screened patients to determine if the ED was the appropriate place. Patients who were assigned to Level 1 were taken directly to the resuscitation room for immediate management. All other patients registered and were formally triaged. Basic investigations were conducted at triage and patients assigned a triage level. ED clinicians assessed patients and investigations requested as needed. Patients were either discharged or referred to inpatient teams. Inpatient clinicians then assessed patients in the ED before making an admission decision.

**Figure 1.**
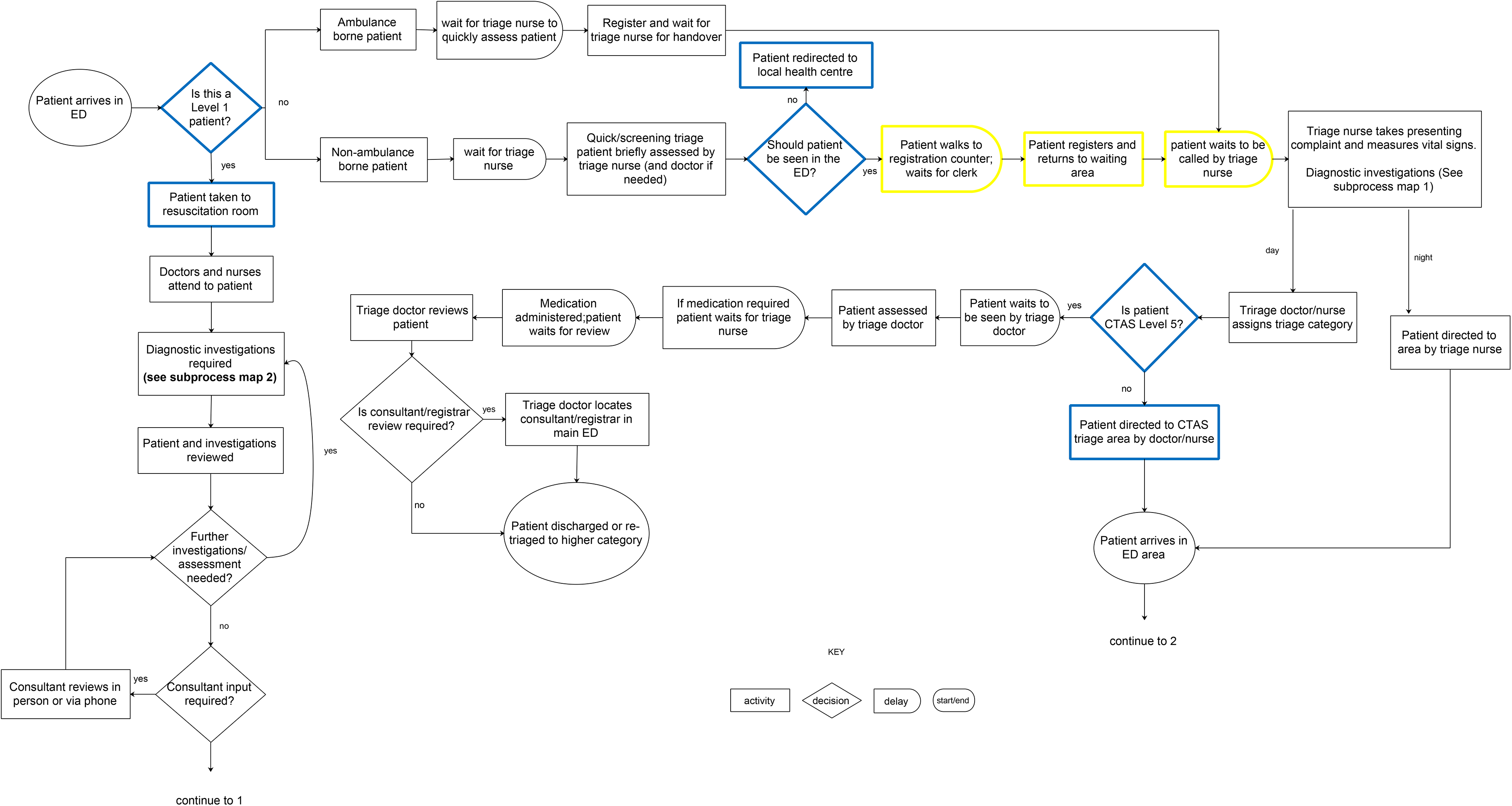
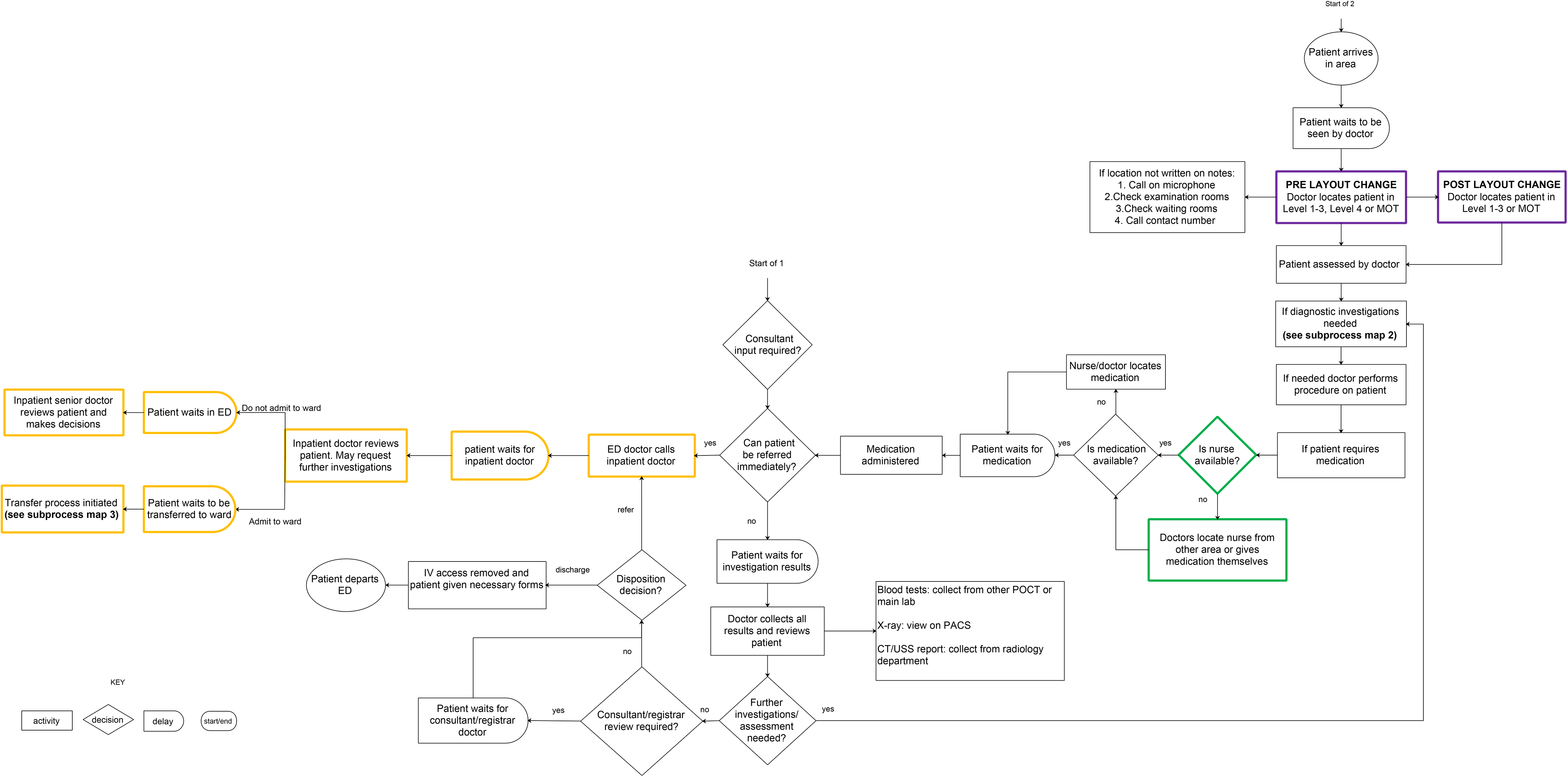
Main process map of patient flow

**Figure 2.**
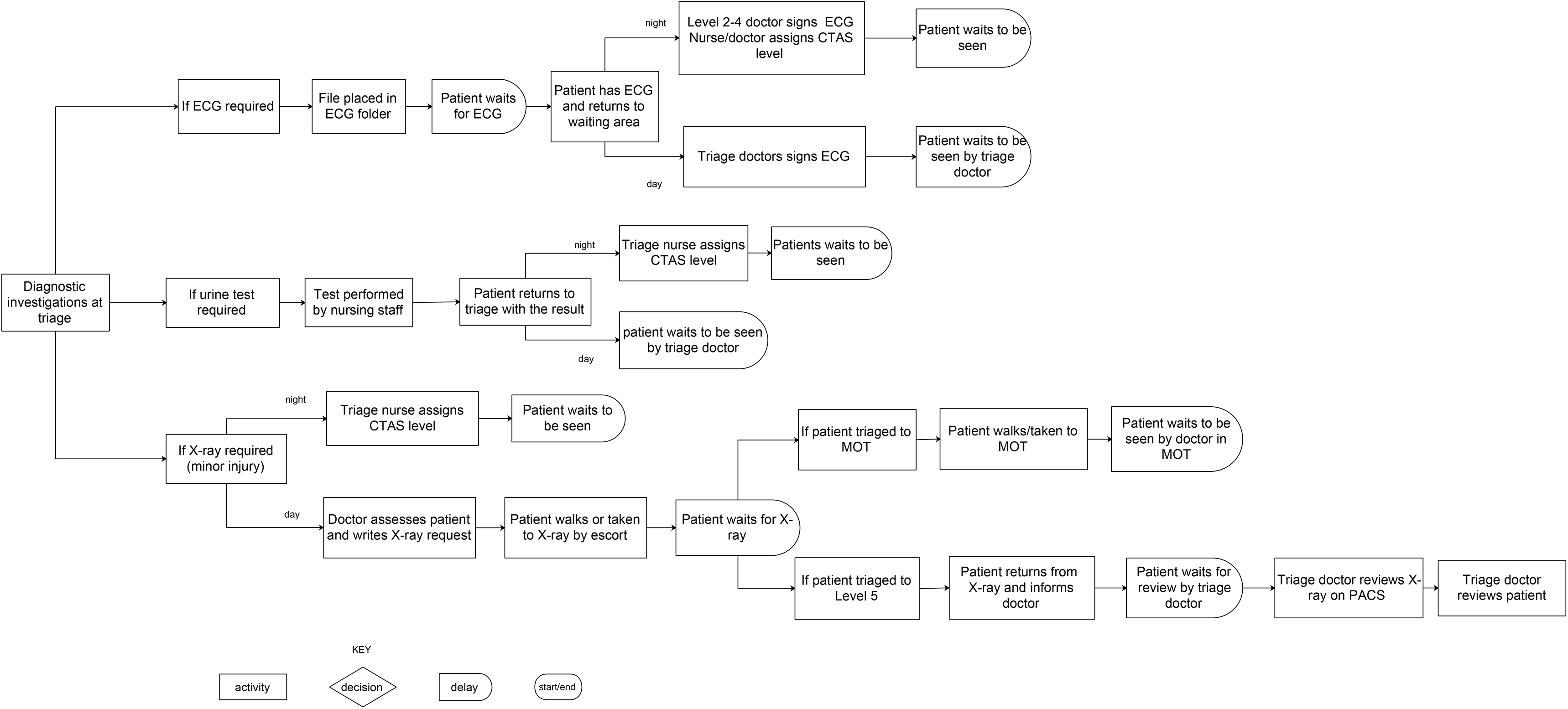
Subprocess map 1-Diagnostic investigations at triage

**Figure 3.**
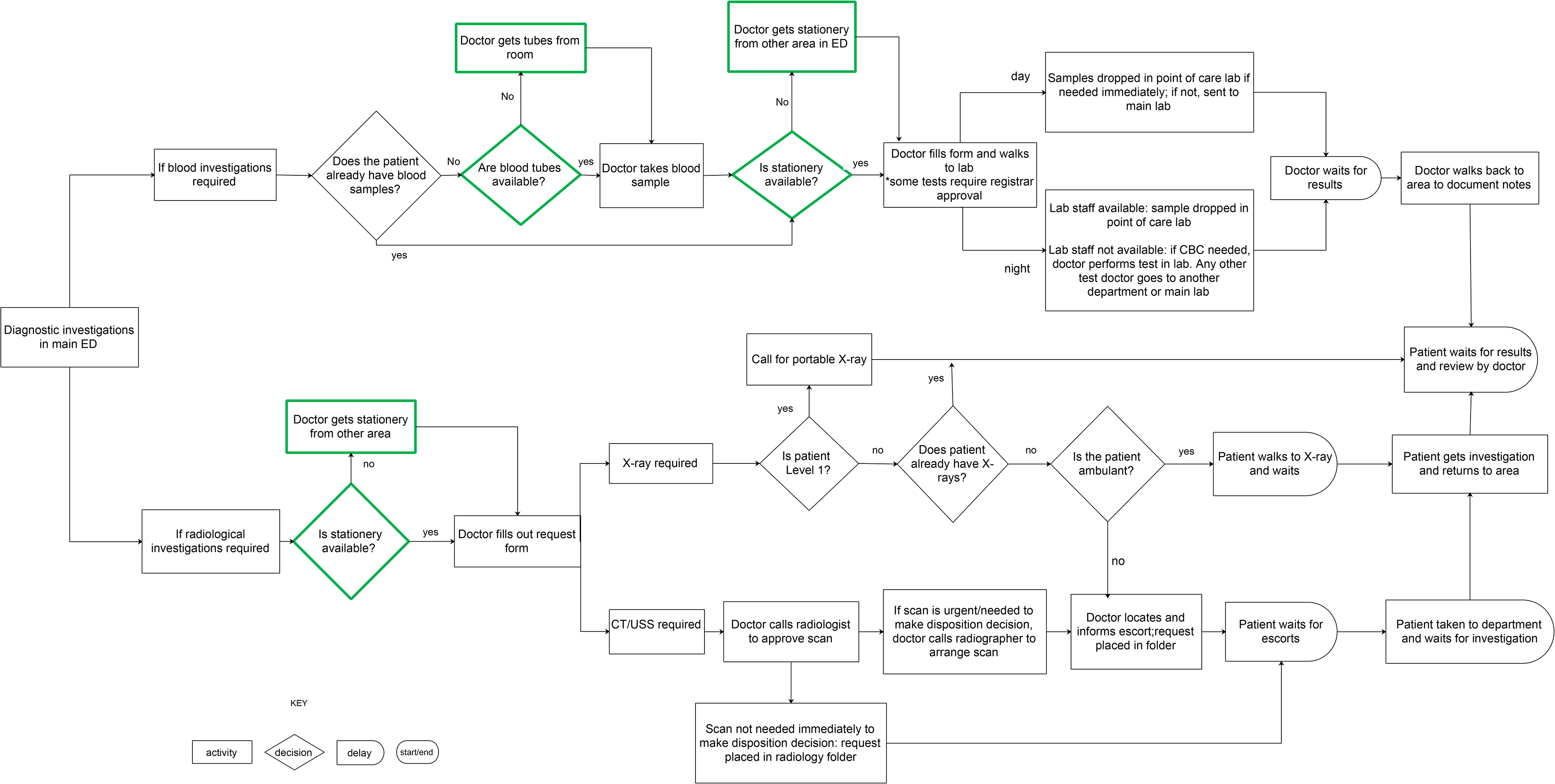
Subprocess map 2- Diagnostic investigations in main ED

**Figure 4.**
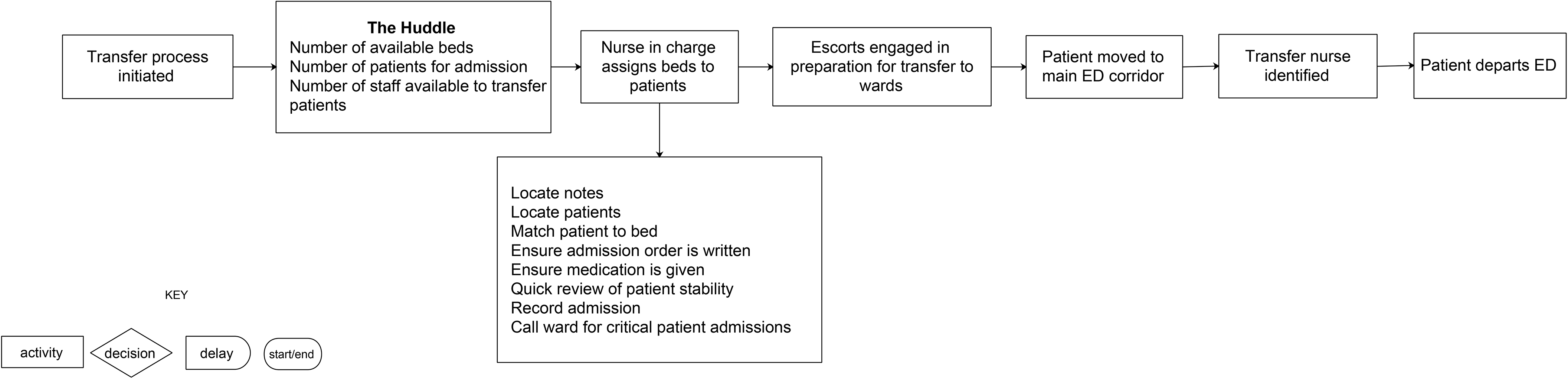
Subprocess map 3- Transfer process

Sub-process maps 1-3 represent key sub-processes related to the patient journey. Sub-process map 1 (figure 2) represents the process for basic investigations conducted at triage. Sub-process map 2 (figure 3) represents the process for diagnostic investigations conducted in the main ED, that is, after patients were assigned to triage categories. The last process map represents the transfer process (figure 4). This map presents the steps taken during the transfer of admitted patients from the ED to inpatient wards.

### ED Reconfiguration

Observations revealed that the reconfiguration was mainly a change in the physical layout of the ED rather than a significant re-arrangement of the steps in the patient flow process. Two main changes were observed: an existing patient examination room that housed non-ambulatory patients was converted to a dedicated examination room for ambulatory patients. The second change was the conversion of the Level 4 area into an area (’holding bay’) to accommodate patients who were either referred to inpatients teams or awaiting admission to the wards. Table 1 Supplementary file 2 summarises the changes in the reconfiguration.

**Table 1.** Summary of ED areas

| ED area | Type of patient seen |
| --- | --- |
| Level 1-3 ('critical area') | CTAS Level 1,2,3 |
| Level 4 | CTAS Level 4 |
| Minor Operating Theatre (MOT) | Minor trauma patients, asthmatics |
| Level 5 ('Triage') | CTAS Level 5, triaging of patients |

### Overarching categories identified as organisational factors influencing the patient flow process

Overall, the analysis generated six overarching categories that appeared to influence patient flow. Within each category there were individual factors that appeared to either facilitate or hinder patient flow. These are presented in the following section with supporting evidence in table 2.

**Table 2.** Organisational factors identified as influencing ED patient flow

| Theme | Subtheme | Evidence |
| --- | --- | --- |
| ED organisational work processes | Facilitator: Streaming<br>Combined with triage<br>Dedicated, space, staff, material resources<br>Staff allocated to each stream<br>Flexible staff redistribution | RN [registered nurse] 2 triaged patient I ... RN 2 took the history while the ENA [enrolled nursing assistant] measured the patient's vital signs. The RN then triaged the patient to Level 5, to be seen by the triage doctor. [Field notes 22, observational process mapping, registration/triage/Level 5]<br>At 2:00pm, a senior [house officer] came to assist. She was actually the doctor assigned to MOT from the 12pm shift but she told me that when she came on shift the critical area was busier than MOT so she went there to assist and clear the area. When that area was under control, she returned to MOT. [Field notes 8, non-participant observations, MOT] |
|  | Facilitator : Front loading of investigations<br>Basic investigations at triage reduced steps in main ED process | This patient presented with chest pain so the triage nurse sent the patient for an ECG. [Field notes 19, observational process mapping, registration, triage, Level 5] |
|  | Facilitator: Flexible assessment options for ambulatory patients<br>Clinically well ambulatory patients assessed on chairs ensuring that a need for trolleys did not delay flow | ...She [team leader] called for patient D over the microphone. He [patient] came walking from the critical [Level 1-3] waiting room. The team leader put him to sit on a chair in the critical area and she assessed him there. [Field notes 16, observational process mapping, Level 1-3 area] |
|  | Barrier :Transfer process | "The first issue is actually locating the notes in the department. The notes are supposed to be placed on the nurses' desk once the patient is for admission. But what can happen is the inpatient teams use the notes while on rounds [in the ED] and they don't return the notes to the nurses. Notes can be left anywhere in the department and occasionally outside the department". [Consultant #2, transcript #3, map review session # 3] |
| ED design and layout | Facilitator: Support organisational work processes | Patient G... sat on a chair near the doctor's workstation. The HO [house officer] took the history then took the patient to BW1 [dedicated examination room] and placed the patient on a bed to examine him. After examining the patient, the patient returned to the chair. [Field notes 33, |
|  | Barrier: Physical separation of areas<br>Barrier: Location of resuscitation room | observational process mapping post layout changes, Level 1-3 area].<br>Patient E walked in via the ambulance bay entrance. The nurse...told the patient to register and then return. The patient walked across to the registration counter, registered then returned to the waiting area. [Field notes 19 observational process mapping, registration/triage/Level 5 area]<br>... For a patient to go from the arrival area to the resuscitation room they would have to pass through the main doors to the interior of the AED down a short corridor then past the HDL[high dependency level] bay potentially navigating patients on gurneys in the corridor. [Field notes 2, non-participant observations, Level 1-3 area]. |
| Material resources | Facilitator: Dedicated ED point of care and radiology<br>Barrier: Insufficient materials created unnecessary motion<br>Facilitator: Staff respond by keeping materials on themselves<br>Barrier: Lack of inpatient beds delayed outflow and increased ED workload | She [house officer] dropped the sample to the POCT [point of care testing] lab and walked back to write her notes... [Field notes 12, observational process mapping, MOT area]<br>There were no more X-ray forms in MOT so he [house officer] walked to the critical area to get a form then walked back to MOT. [Field notes 11, observational process mapping, MOT area]<br>... he [junior house officer] left to get blood bottles from the registrar room...“I fill my pockets with blood bottles so I don’t have to walk back and forth.” [JHO #3, Field notes 24, observational mapping, Level 4]<br>At 1:32am patient E, a patient who was in the ED under the medical team, also crashed [deteriorated]... I asked the HO [house officer] how long patient E had been in the ED and he told me the patient registered at 1:42pm...12 hrs before... the ED team continued to actively resuscitate the patient. [Field notes 18, observational process mapping Level 1-3] |
| ED nursing staff levels, roles, skill mix and use | Barrier: Low nursing staff levels<br>Facilitator: Multitasking and role sharing | There were only 6 nurses on the night shift. They were distributed as follows: 1 nurse assigned to report, 1 to resuscitation, 2 to the critical area, 1 to triage and 1 to share between MOT and Level 4. The nurse sharing between MOT and Level 4 was assigned to the MOT area and was meant to go across to Level 4 if the doctors needed medication. [Field notes 11, observational process mapping, MOT area]<br>I observed escorts moving patients to the main corridor to transfer them to the wards.. a nurse was required to accompany patients but... only two nurses had come to work. [Field notes 27, observational process mapping, Level 4]<br>... on the previous shift there were only four nurses...there were 14 admissions; a nurse was required to go with the transfers but because of the shortage, it was extremely difficult. ...The |
|  | Barrier: Limited nursing roles and skill use<br>Facilitator: Nursing support staff | registrar told me that in the end the nurse in charge had managed to get eight patients transferred – by leaving no nurses in the critical area. She said the nurse in charge left the keys to the dangerous drugs cupboard with her so she could access medication while he and the other nurses transferred the patients. [Field notes 7, non-participant observations]<br>The nurse ... decided that the patient should be triaged to MOT.. he [the nurse] wanted the doctor to review to decide if the patient needed an X-ray so that it could be done before the patient went to MOT. Only the doctor could write the request so the patient had to wait because the triage doctor was assessing other patients. [Field notes 20, observational process mapping, registration/triage/Level 5 area]<br>Patient C came directly to the ENA (enrolled nursing assistant) and complained of chest pain. The ENA sent her directly for an ECG ... When the patient returned the triage nurse took her history while the ENA did her vital signs... [Field notes 19, observational process mapping, registration/triage/Level 5 area]. |
| ED non-clinical staff | Barrier: Task prioritisation affects escort availability<br>Facilitator: Clinical staff adopting escort roles | The house officer ... called the registrar because he needed an escort to carry the patient to CT but the escorts were in the process of transferring patients. He [senior house officer] decided to take the patient across himself rather than wait for the escorts. [Field notes 16, observational process mapping, Level 1-3] |
| ED external clinical and non-clinical departments | Barrier: Inpatient doctors affect outflow<br>Barrier: Reliance on non-clinical departments for reports<br>Facilitator: Clinical staff perform non-clinical roles | ...The patient had been referred to the on-call medical team at 2:15am- the POD [physician on duty] reviewed the patient at 5:45am. [Field notes 18, observational process mapping, Level 1-3]<br>The HO [house officer] decided to request a CT scan for the patient. She called the radiologist at 3:10am to approve the CT... [the patient] went to the radiology department at 3:25am... the patient waited for the CT report, which was not released before I left at 6:00am. [Field notes 26, observational process mapping, Level 4]<br>The team leader was also waiting for a CT report for one of her patients. She told me she was going to walk down to the radiology department to see if any reports were available. [Field notes 16, observational process mapping Level 1-3] |

### ED organizational work processes

The ED organizational work processes relevant to patient flow were identified as streaming of patients, front loading of investigations, flexible assessment options for ambulatory patients and the transfer process. These processes were implicit or intuitive strategies observed/ recounted rather than explicitly documented policies in the department.

#### Streaming, allocation and re-distribution of staff facilitates simultaneous flow of multiple patient groups

The combined streaming and triage process appeared to facilitate flow, prioritising seriously ill patients at the onset of the patient journey. Each stream had its own dedicated space, staff and material resources allowing staff to simultaneously assess multiple patient groups. The process map in figure 1 highlights the decision and activity steps that reflect the streaming process (steps marked blue). The allocation of clinical staff to each stream also facilitated patient flow. Doctors (house officers) and nurses were assigned to each stream with greater numbers of clinical staff assigned to higher priority streams. Lastly, there was flexible redistribution of staff to match areas of demand. The combination of these factors appeared to promote good patient flow.

#### Frontloading of investigations at triage reduced steps for patients

The front loading of investigations intended to facilitate patient flow. Requesting basic investigations (ECGs, urine tests, X-rays for minor injuries) during the triage process appeared to improve flow by reducing the number of steps after the main clinical assessment. Figure 2 presents the process map of the front loading of investigations during the triage process.

#### Flexible assessment options facilitated flow for ambulatory patients

Observations revealed that patients were not automatically placed on trolleys in order to be seen by doctors. Doctors identified reasons such as patients being well enough to sit, insufficient trolleys and the need to anticipate future patients who may require a trolley, illustrated in the following extracts.

> “No, everyone can’t get a bed because there aren’t enough and even if there were available beds we wouldn’t put someone on a bed if they didn’t really need it. You also have to anticipate that someone else may come in who really needs the bed.”[Registrar #8, nonparticipant observations]

Clinically well ambulatory patients were often seen on chairs. This strategy of utilizing chairs to assess patients was not a formal policy in the ED but appeared to be an implicit strategy aimed at prioritising trolleys for patients most in need. As a result, staff often spent time searching for available space to use. Overall, the strategy itself appeared to facilitate patient flow since ambulatory patients did not have to wait for an available trolley to be seen and supported the appropriate utilization of trolleys.

#### Transfer process delays the outflow of admitted patients

The transfer process referred to the movement of admitted patients from the emergency department to inpatient wards (figure 3). This was a complicated sub-process with multiple factors affecting each step with some factors facilitating outflow and others acting as barriers to good outflow. One aspect intending to facilitate patient outflow was a team meeting, (‘the huddle’), that occurred at several intervals throughout the day. ED staff were regularly updated on the numbers of available inpatient beds, patients for admission and staff available to assist with patient transfers. This strategy was thought to provide ‘structure and coordination’ to the transfer process [Consultant#2].

Other observed factors appeared to act as barriers to the outflow of admitted patients. The activity of assigning admitted patients to inpatient beds comprised multiple steps, which appeared to consume staff time. Locating patient files was time consuming because of the involvement of external clinical staff who often did not return files to the nursing staff. Locating patients in the department was also a barrier because the patient location was not always documented on the files. Further delays in the process resulted from a lack of nurses and attendants required to transfer the patient.

### ED design and layout

#### ED design and layout facilitated flow by supporting the organizational work processes

The ED layout appeared to support the streaming process by having distinct separate areas for each stream (supplementary file 1). The physical reconfiguration also highlighted the influence of design on patient flow. The introduction of an examination room specifically for ambulatory patients appeared to support the flexible assessment organizational work process and reduced time staff spent searching for available space.

#### Features of the ED layout created additional steps in the process

Layout features that appeared to hinder flow included the physical separation of the registration and triage areas. The separation of these areas created additional activity and waiting steps in the process which are reflected in the highlighted yellow steps in figure 1.

In the physical reconfiguration, dedicated ED areas for referred or admitted patients (holding bays) were also introduced. This appeared to be useful for the overall organization of the ED by separating admitted patients from those still receiving emergency care but overall, it appeared that the reconfiguration did not substantially alter the steps in the patient flow process. The process map (figure 1 highlighted purple steps) showed that the patients experienced the same steps but in a different area within the ED.

### Material resources

#### Dedicated ED laboratory and radiology services facilitated patient flow

Dedicated ED point of care testing and X-ray services appeared to facilitate flow by providing results in a timely manner and reducing dependency on external departments.

#### Insufficient material resources in the ED led to increased motion searching for materials

Insufficient materials, such as phlebotomy and stationery materials, created unnecessary motion from staff searching for materials acting as a barrier to flow. The highlighted green steps in sub-process map 2 (figure 4) show how insufficient materials in the ED created additional steps in the process. Subsequent observations revealed that staff responded to the insufficiency by keeping specific materials on themselves to reduce time spent searching.

#### Lack of inpatient beds appeared to be a barrier to the outflow of admitted patients (transfer process)

Staff also noted the lack of available inpatient beds as a factor affecting outflow with one staff member stating, “The biggest bottleneck in transferring patients out of the department is the lack of beds on the ward…”[Head nurse#1]. Further observations showed that this led to patients boarding in the ED which increased the workload for ED staff and exacerbated other factors influencing patient flow such as the shortage of nursing staff, described in the next theme.

### ED nursing staff levels, roles, skill mix and use

#### Nursing shortages compromised nurse dependent steps leading to sharing of roles amongst staffing groups

Observations and field conversations revealed that each shift required fourteen nurses but this number was not always met. The nursing shortage appeared to be most significant on night shifts, affecting the allocation of nurses to ED areas, leaving some areas unstaffed, which consequently acted as a barrier to effective streaming. The nursing shortage also led to delays in the triage process, administration of medication and the transfer of patients out of the ED. Highlighted green sections of Figure 1 show how the nursing shortage delayed administration of medication and created extra steps in the patient process.

The nursing shortage resulted in nursing staff and doctors adjusting their roles to meet the demands of the department. Observations revealed that nurses multitasked, often assigned to manage multiple streams and doctors shared nursing roles to counter shortages. For example, in one instance a doctor shared nursing duties to allow the nurses to complete the transfer process.

#### Limited nursing roles and skill use created more doctor dependent process steps

Observations revealed that nurses were unable to institute patient management, perform invasive clinical procedures or request investigations. Limited nursing roles appeared to influence the effectiveness of work processes, such as front loading of investigations, since only doctors could authorise requests for investigations. Registered nurses with additional training were not always able to utilize their skills because they mainly performed administrative roles. However, the nursing shortage affected nursing skill use, as one head nurse explained:

> “Even if nurses were allowed to do more, the current numbers wouldn’t allow them to see patients because it would take away from the general nursing care required” [Head nurse#2,]

Lastly, within the overall nursing staff category, there were a variety of auxiliary staff who supported registered nurses in their nursing duties, promoting flow.

### ED non-clinical staff

#### Multiple duties of escorts affected escort availability acting as a barrier to patient flow

Patient progression often depended on availability of the escort staffing group. There was often conflict regarding which task (patient transfers to wards or transporting patients for investigations) should be prioritised. Although these duties facilitated flow for one group of patients it hindered flow for the other group. Similar to the response to the nursing shortage, doctors carried out tasks that escorts would normally be expected to undertake, in order to maintain flow.

### External clinical staff and non-clinical departments

#### Dependency on external departments delayed decision-making and patient outflow

Observations showed that external clinical staff, that is, non-ED doctors, appeared to influence flow, acting as a barrier to patient outflow. When patients were referred to inpatient doctors, these doctors assessed the patient in the ED before making their disposition decision. This often involved clinical assessment (history and examination) and requesting of further investigations. ED staff considered the rate at which the inpatient doctors assessed patients a major obstacle to patient flow.

> “This is the biggest delay in the department-waiting for the specialty teams to review the patient” [SHO#16].

As seen in Figure 1 (highlighted orange steps), the inpatient team influenced the steps taken after an ED disposition decision was made.

Delays in receiving reports from non-clinical departments, such as the main hospital laboratory and radiology departments, appeared to influence flow not only because of longer waiting times but also because of a lack of a mechanism to alert doctors when results were ready. Again, doctors opted to perform non-clinical tasks, such as walking to departments to collect reports.

### Conceptual model of factors influencing patient flow

The findings from the literature review and the primary study were summarised in a conceptual model of factors influencing ED patient flow (figure 5). The model builds on the existing qualitative literature by providing further insight and explanation into how identified factors influenced patient flow. In the model, the findings were re-organized into six categories, based on a modified fishbone model [20]. Within the categories, the model identifies specific factors that are considered either barriers or facilitators to patient flow. Although the model classifies the factors into broad categories, these factors do not exist in isolation. For example, while streaming and triage (Methods) created simultaneous pathways and was considered a facilitator of patient flow, the method is dependent on having sufficient staff (Staffing) to allocate to each stream (People). Thus, the model summarises the findings on the factors influencing ED flow and provides a structured approach to understanding patient flow.

**Figure 5.**
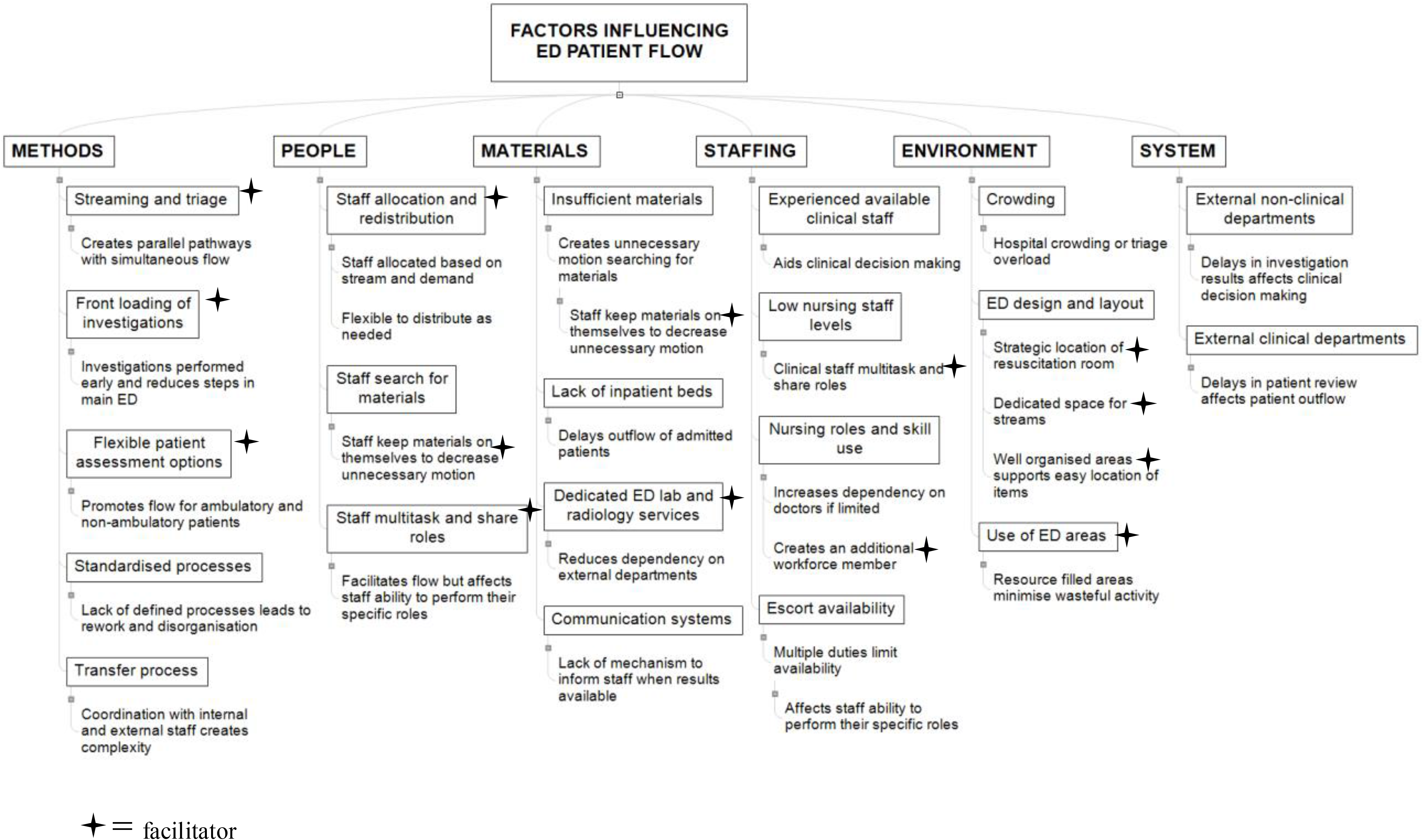
Conceptual model of factors influencing ED patient flow

## DISCUSSION

This study used qualitative methods, primarily observational process mapping, to explore patient flow in an ED in a developing Caribbean island. The findings in the study are consistent with existing literature from both developed and developing countries. Factors common to other studies included a lack of inpatient beds and material resources, staff shortages and impact of inpatient teams [21-27]. The transfer process in the primary study required detailed coordination and cooperation within and outside the ED. This was consistent with an American study that described similar challenges with the outflow of admitted patients from the ED to inpatient wards [21].

In the primary study, clinically well ambulatory patients were assessed on chairs, facilitating flow for this patient group. This is similar to a ‘fit to sit’ strategy in the UK where suitable ambulance borne patients were placed on chairs on arrival to the ED [1].The strategy also has similar characteristics to flexible care areas or rapid assessment zones described in studies conducted in developed countries [28,29]. These strategies involved the use of dedicated spaces to treat patients for whom trolleys were not considered necessary. However, in the current study, there were no formally documented departmental policies for any of the identified organizational work processes. For example, there were no criteria detailing which patients were appropriate for the flexible assessment option. These strategies may be generalisable to other settings (or may already exist in some form, as in the primary study) but standardization of the intervention, formalizing policies reduces guesswork and unnecessary activity, ultimately supporting good patient flow.

The ED reconfiguration undertaken in the primary study also highlighted the influence of design on patient flow, supporting the suggestion that design strategies should facilitate (effective) work processes while also demonstrating the importance of considering how movement and activities of process users affect flow [30]. Using this approach should aid decision makers when determining if restructuring the ED is a viable strategy to address flow concerns.

The nursing shortage and the limited use of nursing skills identified in the primary study, was also a factor affecting flow in other emergency departments [22-24, 31]. Nursing shortages are common in EDs regardless of the setting [43]. However, nursing levels in developing countries are often further compromised because of migration from developing to developed countries [32]. The UK Royal College of Nursing states that safe and effective staffing means ‘having enough nursing staff with the right skills and knowledge, in the right place, at the right time’ [33]. Based on this, the ED case study had low safe nursing staff levels. Nursing roles such as emergency nurse practitioners are established in developed countries but are less common in developing countries [34]. These are likely to be valuable in developing settings but require legislation, education and professional support for proper implementation [34].

Staff actions such as multitasking and role sharing were often in response to increasing demands in the ED or perceived barriers to patient flow. This behaviour was noted in other studies with staff manipulating ED space by re-distributing patients to areas that were less busy or by staff persistently calling the external departments to remind them about the reports for investigations [21, 22, 35] However, while these actions may have facilitated flow, if they become sustained or permanent, it may affect the staff ability to perform their primary roles, which subsequently hinders patient flow.

### Strengths and Limitations

There are several limitations to this study. The data collection occurred in a single ED in Trinidad, which may not reflect the processes in all EDs in the country or other settings.

Future research should focus on conducting larger studies across a wider range of settings to validate the findings. The fieldwork was also conducted by a single observer which may lead to researcher bias. However, several methods were used to minimize this risk. These included a prolonged length of time in the field, triangulation of data using multiple methods and data sources, sharing of transcripts with other authors and validation of process maps with key staff members.

Time constraints limited the number of hours of observations on admitted patients who remained in the ED, which meant that this stage of the patient journey was not completely explored. The limited use of verbatim speech in the informal conversations may have affected the reliability of this data. Additionally, participants may have adjusted their behaviour in response to the observer’s presence. However, the length of time in the field, the nature of the ED being an intense environment with staff who are likely to be constantly occupied and the high patient turnover, may have reduced this effect. This study also did not explore areas such as organizational culture, professional relationships or power imbalances, which may provide additional insights into patient flow. Future studies, in addition to exploring the organisational patient flow process, may also benefit from incorporating how these areas influence patient flow and the organisational process.

In conclusion, this study contributes to knowledge on emergency care research in the Caribbean and may be relevant to other developing countries. The findings may be a step towards strengthening the ED in the local context, supporting the WHO emergency care systems objectives. The study findings also suggest that there are common flow concerns across settings; combining efforts has the potential to produce robust solutions. However, future research is needed to validate the study findings using larger studies across a wider range of settings.

## Data Availability

The data is in the form of confidential observations and interviews and is not available for data sharing

## Other Statements

### Author statement

LD, SG, ROH, PT conceived the study. LD executed the study including data collection, analysis and management. SG, ROH and PT supervised LD (doctoral student) and provided advice on the study design, analysis and interpretation of results. SH acted as a local supervisor for LD during the data collection period at the case study site. LD drafted the article and all authors contributed to the revision of the article. LD takes responsibility for the paper as a whole.

## Acknowledgments

sincere gratitude to the staff and patients at the Eric Williams Medical Sciences Complex Emergency Department

